# Profiling health and socioeconomic disadvantage in the northern Adelaide Local Health Network population

**DOI:** 10.1101/2024.01.02.24300748

**Authors:** Cheryl Shoubridge, John Maddison, John Lynch, Mark Boyd

## Abstract

**Objective:** This study aimed to describe and compare aspects of the socioeconomic and health status of the population within regions of an Australian capital city.

**Design:** This comparative study harnessed publicly available, deidentified, population level data from the 2021 census spanning measures of relative disadvantage, population demographics, health status and risk factors known to contribute to health outcomes. We describe data for greater Australian capital cities and compare aggregate public health area data that reflect local health network boundaries within greater Adelaide.

**Results:** Northern Adelaide is a region of greater relative disadvantage, demonstrated by the low Index of Relative Socio-Economic Disadvantage score (945) compared to the national average (1000) and scores for the central and southern Adelaide regions. Social determinants that contribute to this relative disadvantage include the proportion of people with no or limited education (26%) and those living on constrained income sources (e.g. unemployment benefits / aged pension, 10% and 72%, respectively). The northern Adelaide region has higher burdens of long-term health conditions including but not limited to diabetes, heart disease, kidney disease and lung diseases. We demonstrate that the comparatively high prevalence of obesity (37%) for people in the north of Adelaide were correlated with low numbers of people with adequate fruit intake (48%) and the higher proportion of people who currently smoke (16%) and who undertake low or no levels of exercise (73%).

**Conclusions:** Social disadvantage in the northern Adelaide region compared to the less disadvantaged central and southern regions of Adelaide is associated with poorer health outcomes and correlated with higher levels of health risk behaviours. Understanding the challenges within this local community setting may provide opportunities for local health networks to lead interventions to mitigate social risks and thereby improve health outcomes in this population.

**Short Key Question Summary:** *What is known about the topic?:* Social disadvantage is correlated with poor health outcomes

*What does this paper add?:* Profiles the types and prevalence of social determinants of health and the health status of a community region within an Australian capital city.

*What are the implications for practitioners?:* This profile of adverse social determinants of health relevant to the local community setting provides a context for the health system to respond, advocate for and intervene effectively to improve the well-being needs of this disadvantaged community.

## Introduction

Social determinants of health (SDoH) are the ‘non-medical factors influencing health outcomes’ and can both enable and promote, as well as create barriers to achieving optimal health outcomes ^1^. SDoH are a broad category of factors that include but are not limited to housing, neighbourhood and physical environment, safety, food availability and financial security. When communities and populations experience social disadvantage there is a well-established connection with adverse health outcomes, underscored by the 2008 WHO Commission on Social Determinants of Health report ^2^. Comparison of health risk factors, chronic conditions, deaths and disease burden in different socioeconomic groups in the Australian setting demonstrate that across almost all health measures, people living in lower socioeconomic groups generally have worse health outcomes, and in 2018 over one-third of disease burden was potentially preventable^3^. This phenomenon is known as ‘the social gradient of health’. It is estimated that the relative contributions to health and wellbeing are ∼20% for the health system and ∼50% by SDoH ^4-6^. The profile of socioeconomic disadvantage not only contributes substantially to increased disease burden but may also constitute barriers to effective treatment and intervention.

The greater Adelaide region is serviced by three metropolitan local health networks (LHN), comprising the northern (NALHN), central (CALHN) and southern Adelaide (SALHN) local health networks. The northern Adelaide community has long been recognised as disadvantaged and is well understood to manifest relatively poor health outcomes ^4^. This population is served by NALHN, comprising two major hospitals, the Lyell McEwin and Modbury Hospitals. We contend that profiling the health status and contributors to socioeconomic disadvantage within this northern Adelaide population may identify opportunities for local health network to lead social interventions to improve the health of this disadvantaged community.

## Materials and Methods

### Data source

We analysed data from the Australian Bureau of Statistics (ABS), using the Social Health Atlas of Australia: 2021 census and data by population health areas (PHA)^7^, updated throughout 2023. The Index of Relative Socio-Economic Disadvantage (IRSD) score developed by the ABS as part of the Socio-Economic Indexes for Areas (SEIFA) is standardised to the national average of 1000 and standard deviation of 100, with a score <1000 indicating a relatively greater disadvantage. SEIFA data was sourced from (https://phidu.torrens.edu.au). PHA codes and names from the South Australian data for the greater Adelaide region, and the catchment areas for the individual LHNs are shown in Supplementary Information -Table 1. Ethics approval was not required for this study as only publicly available, deidentified, population level data was used.

**Table 1:**
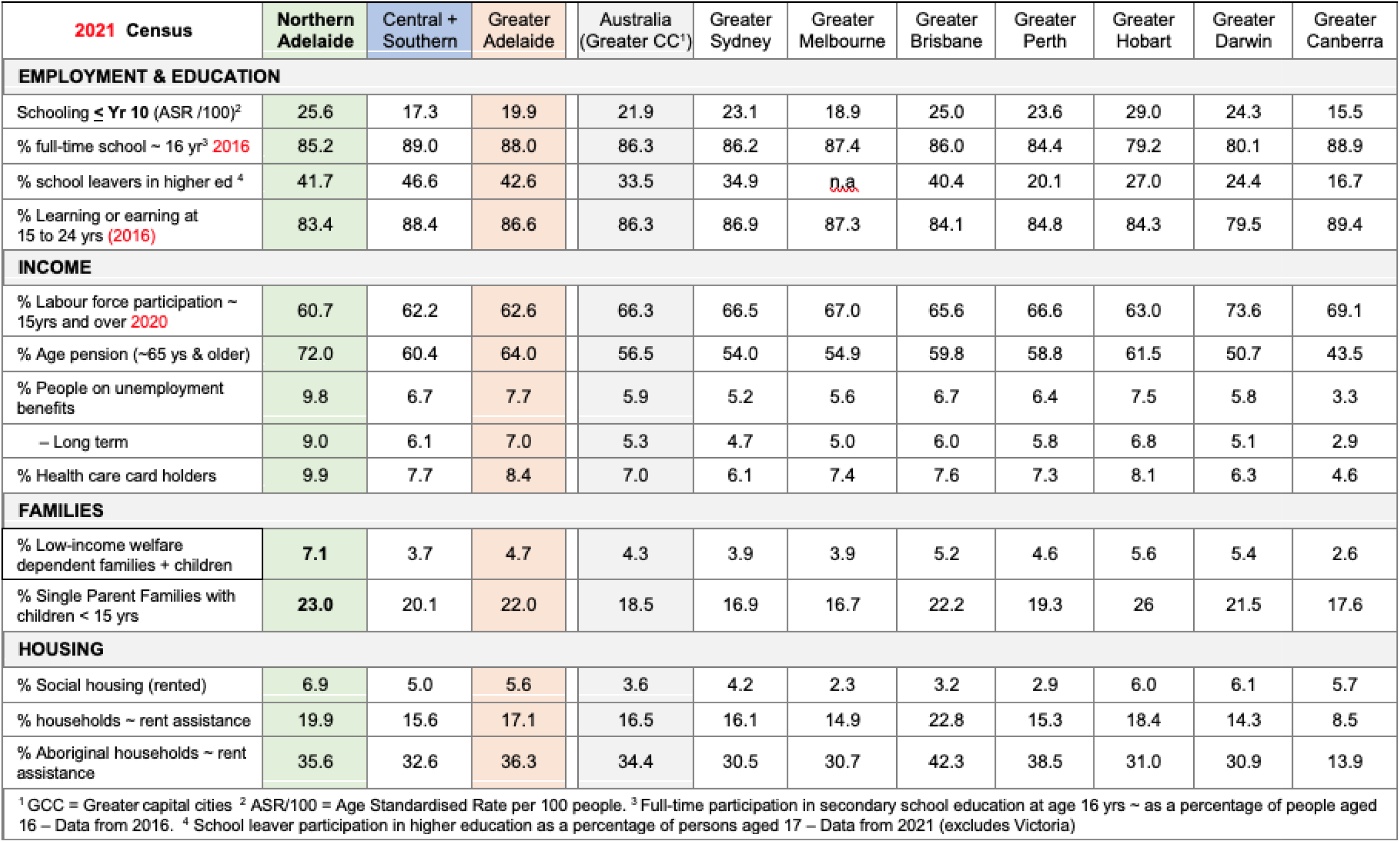
Factors contributing to disadvantage in the greater capital cities and greater Adelaide region.

### Data and statistical analysis

Data were analysed using GraphPad Prism version 9 (GraphPad Software Inc.). For data presented as box and whisker plots, the median, first, and third quartiles were calculated for each group, with whisker boundaries representing the minimum and maximum values. Pairwise comparisons were analysed using simple linear regression.

## RESULTS

### The northern Adelaide region is a region of disadvantage

The Index of Relative Socio-Economic Disadvantage (IRSD) scores from the 2021 census for greater capital city areas within each state of Australia demonstrate that greater Adelaide and greater Hobart have scores below the national average, with 993 and 991, respectively (Figure 1A). The IRSD score in the northern region (945) indicates greater disadvantage compared to the scores in the central (1024) or southern region (1007) of greater Adelaide at the local government area (Figure 1B). Thirteen PHAs within the local government areas of the northern region have IRSD scores below the national average of 1000 (Figure 1C). Of the 18 PHAs within the northern Adelaide region, 6 were ranked within the worse ∼10% disadvantaged regions nationally (Figure 1D).

**Figure 1:**
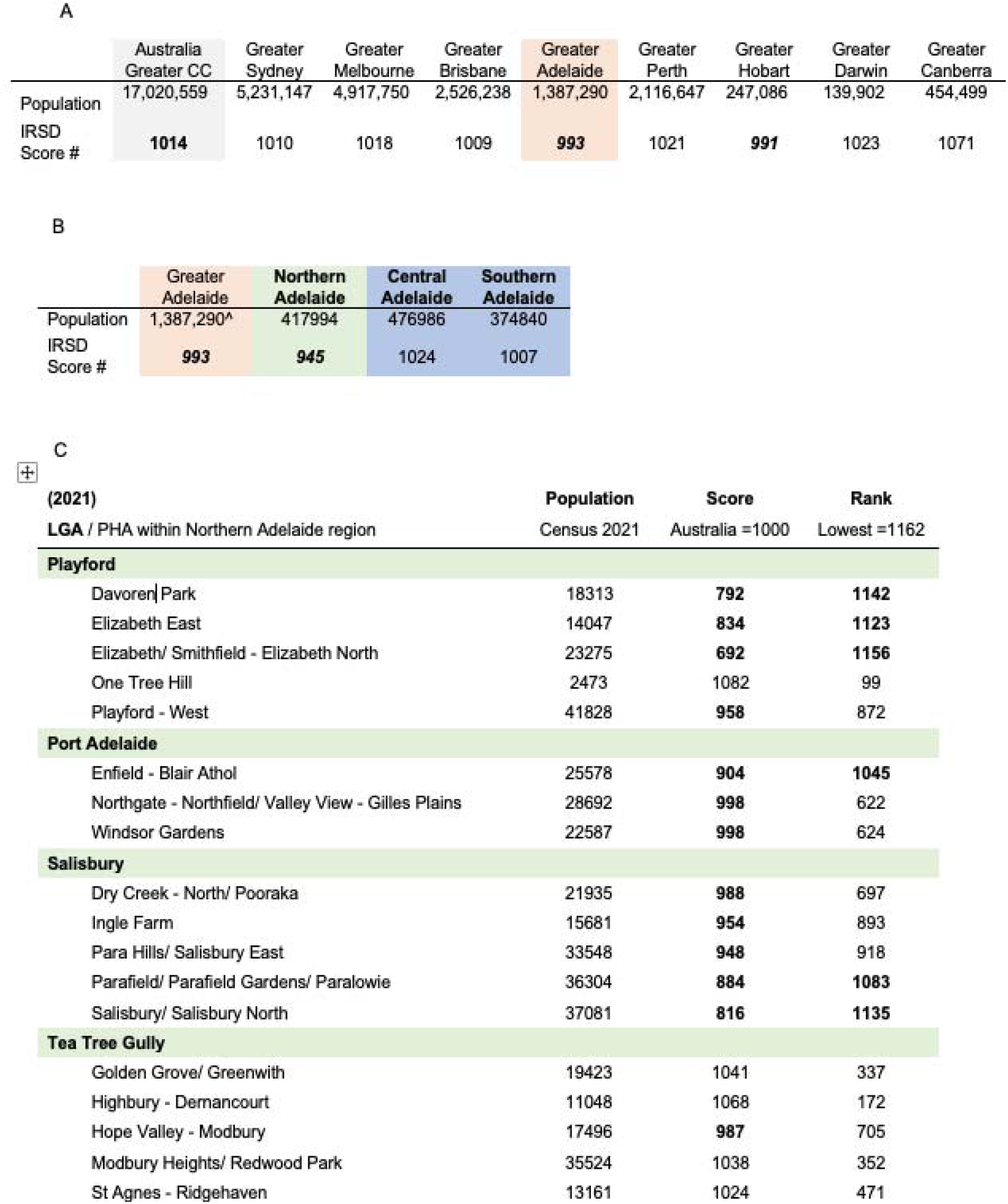
IRSD scores in greater Adelaide is being driven by the relative disadvantage in the northern Adelaide region. Population and IRSD scores for the A) greater capital cities and B) LHNs (^numbers of people in greater Adelaide Area are higher than the aggregate values of southern, central and northern regions due to a small number of PHAs that are captured in the greater Adelaide region that fall outside the catchment PHAs for the three metropolitan Local Health Networks within Adelaide). C) National ranking of IRSD scores indicate that 13 LGAs within northern region have lower scores than the national average (bold), with 6 PHAs (bold) in this region ranked within the 10% most disadvantaged. All data from the 2021 census.

### Population composition

The northern Adelaide region comprises ∼ 24% of the greater Adelaide population with residents born in Australia (64%), overseas in English-speaking (8.2%) or non-English speaking countries (21.3%), consistent with the Australian greater capital cities (61.3%, 8.5% and 25.8%) and the greater Adelaide region (68.7%, 8.4% and 19.2%), respectively (Figure 2A). The proportion of Aboriginal and Torres Strait Islander residents in the northern Adelaide region (2.4%) was higher than those in the central and southern Adelaide region, but less than some other national jurisdictions (Figure 2B and Supplementary Table 2). Northern Adelaide has the equivalent to 44% of the greater Adelaide Aboriginal population and 24% of the total South Australian Aboriginal population (Figure 2C). In terms of age, the largest proportion of people is distributed in the 25 to 44 yr bracket consistently across populations of greater capital cities of Australia (Supplementary Figure 2). In the northern Adelaide region, there were more (60%) young people (under 44 years of age as a percentage of total persons in each region) compared to (55%) central and southern regions of greater Adelaide (Figure 2D).

**Table 2:**
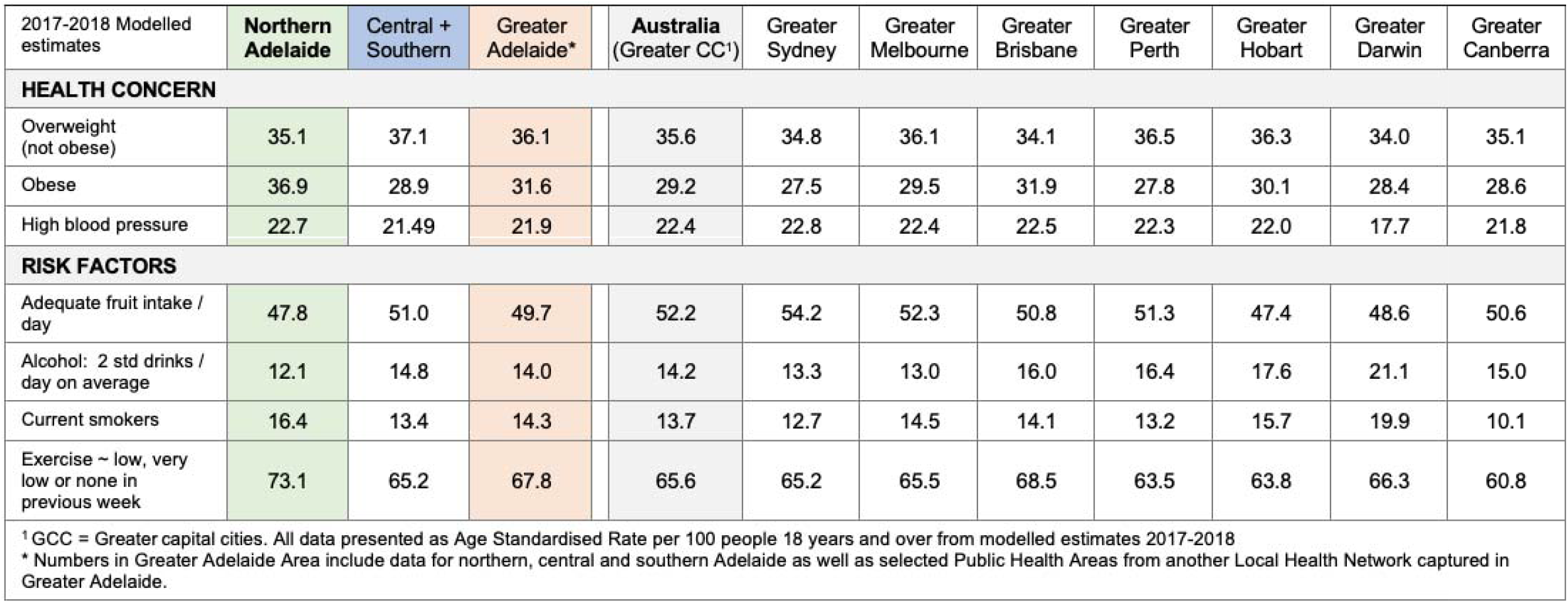
Estimated prevalence of risk factors impacting health in greater capital cities and in Adelaide LHNs.

| 2017-2018 Modelled estimates | Northern Adelaide | Central + Southern | Greater Adelaide* | Australia (Greater CC<sup>1</sup>) | Greater Sydney | Greater Melbourne | Greater Brisbane | Greater Perth | Greater Hobart | Greater Darwin | Greater Canberra |
| --- | --- | --- | --- | --- | --- | --- | --- | --- | --- | --- | --- |
| **HEALTH CONCERN** |  |  |  |  |  |  |  |  |  |  |  |
| Overweight (not obese) | 35.1 | 37.1 | 36.1 | 35.6 | 34.8 | 36.1 | 34.1 | 36.5 | 36.3 | 34.0 | 35.1 |
| Obese | 36.9 | 28.9 | 31.6 | 29.2 | 27.5 | 29.5 | 31.9 | 27.8 | 30.1 | 28.4 | 28.6 |
| High blood pressure | 22.7 | 21.49 | 21.9 | 22.4 | 22.8 | 22.4 | 22.5 | 22.3 | 22.0 | 17.7 | 21.8 |
| **RISK FACTORS** |  |  |  |  |  |  |  |  |  |  |  |
| Adequate fruit intake / day | 47.8 | 51.0 | 49.7 | 52.2 | 54.2 | 52.3 | 50.8 | 51.3 | 47.4 | 48.6 | 50.6 |
| Alcohol: 2 std drinks / day on average | 12.1 | 14.8 | 14.0 | 14.2 | 13.3 | 13.0 | 16.0 | 16.4 | 17.6 | 21.1 | 15.0 |
| Current smokers | 16.4 | 13.4 | 14.3 | 13.7 | 12.7 | 14.5 | 14.1 | 13.2 | 15.7 | 19.9 | 10.1 |
| Exercise ~ low, very low or none in previous week | 73.1 | 65.2 | 67.8 | 65.6 | 65.2 | 65.5 | 68.5 | 63.5 | 63.8 | 66.3 | 60.8 |

<sup>1</sup> GCC = Greater capital cities. All data presented as Age Standardised Rate per 100 people 18 years and over from modelled estimates 2017-2018
\* Numbers in Greater Adelaide Area include data for northern, central and southern Adelaide as well as selected Public Health Areas from another Local Health Network captured in Greater Adelaide.

**Figure 2:**
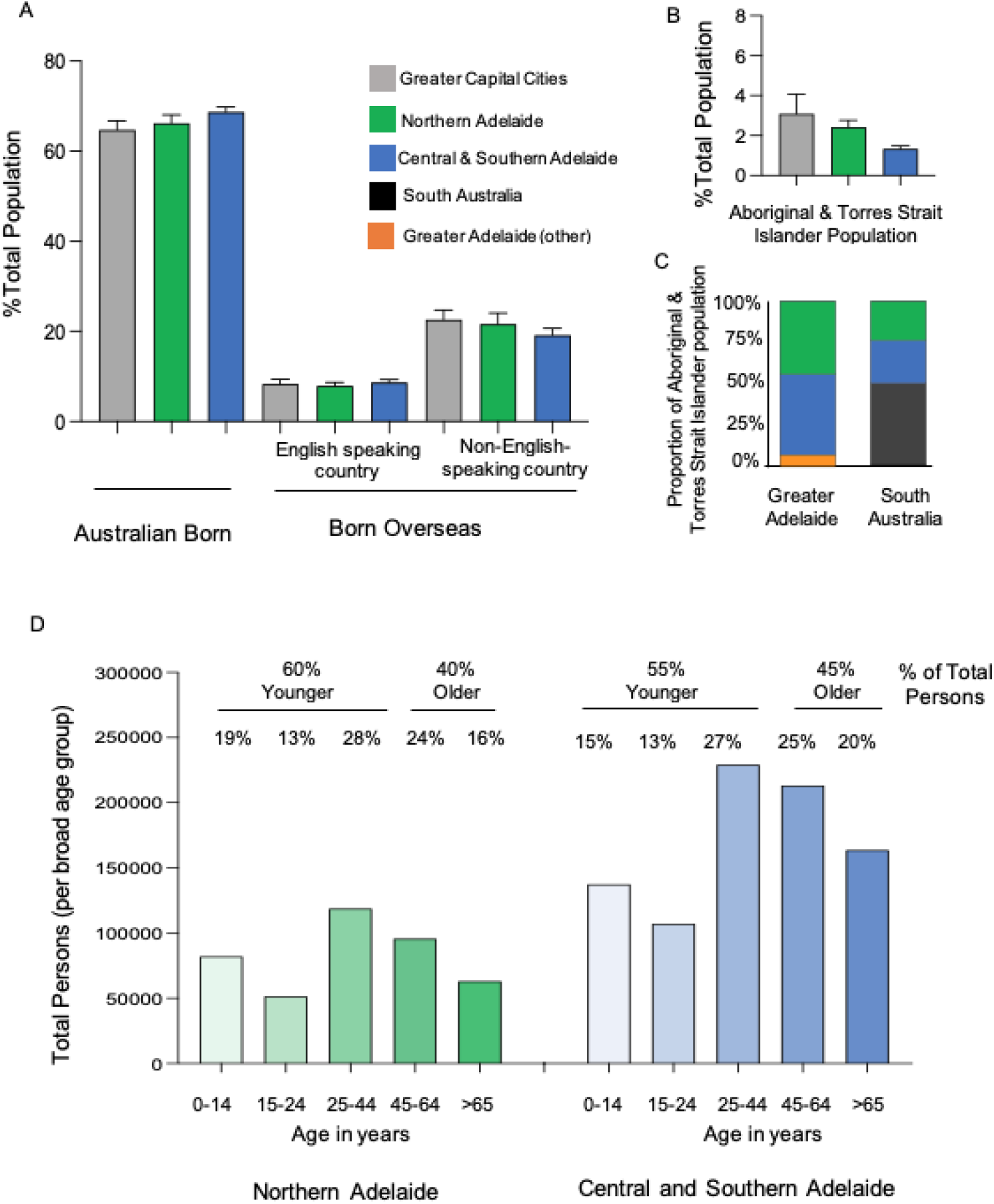
Population country of origin in northern Adelaide is generally consistent with the greater Capital cities and greater Adelaide population landscape. A) Place of birth as a percentage of the total population with greater capital cities and of population health areas within the greater Adelaide regions. People that identify as Aboriginal or Torres Strait Islander as a B) percentage of the total population Greater capital cities (grey bars), northern Adelaide region (green bars) and central and southern Adelaide regions (blue bars). Data presented as Mean ± SEM. C) People that identify as Aboriginal or Torres Strait Islander as a as a proportion of the greater Adelaide (left) and South Australian population (right). Northern Adelaide region (green bars) and central and southern Adelaide regions (blue bars), remaining Greater Adelaide region outside northern, central and southern regions catchments (orange bars) and remaining South Australian population (black). Data from the 2021 census. D) Age distribution for northern region (green bars) and central and southern regions combined (blue bars) (Total persons). The percentage of younger persons (under 44 years) and older persons (above 45 years) are indicated above the individual bars. Data from 2020 estimated resident population released in 2021.

### Social Determinants: education, income, families, and housing

The northern Adelaide region had one of the highest rates (at 26 per 100) of people who did not go to school or left school at or below Year 10 compared to other greater capital cities and other regions of greater Adelaide (at 17 per 100) (Table 1). For the working age (16-64 yrs) and aged population (65 yrs and over), the greater Adelaide region (7% and 64%) nationally and the northern Adelaide region (10% and 72%) locally have the highest percentages of people receiving unemployment benefits (many as long-term recipients ∼greater than 6 months) and the aged pension (Table 1). The proportion of low-income welfare dependent families with children in the northern Adelaide region (7%) is higher than all the greater capital cities (median at 4.5%) (Table 1). The percentage of social housing (rented dwellings) and households receiving rent assistance is variable across the greater capital cities of Australia with a higher proportion in the northern compared to the combined central and southern regions of Adelaide for both measures (Table 1). The distribution of these measures for individual public health areas (PHA) are shown as individual points on the graphs in Supplementary Figure 3.

### Long-term health conditions, health measures and hospital admissions

The number and types of long-term health conditions, median age of death and the prevalence of premature mortality and avoidable deaths are profiled on Supplementary Table 3. Relative to the greater capital cities of Australia, greater Adelaide has the third highest level of premature mortality (236 per 100,000), with the northern Adelaide region (271 per 100,000) higher than central and southern Adelaide (224 per 100,000). This would equate to 1132 people dying prematurely in the northern Adelaide region, 197 more deaths relative to the deaths in the central and southern regions of greater Adelaide for a comparable population across a similar time frame. Similarly, death from avoidable causes were higher in northern Adelaide region (at 131 per 100,000) (Figure 3A) compared to the greater Adelaide region (at 111 in 100,000) (2016 to 2020 in person 0-74 years - deaths from conditions that are potentially preventable through individualised care and/or treatable through existing primary or hospital care - Supplementary File 1). This would equate to 105 more avoidable deaths in the northern Adelaide region relative to other regions of greater Adelaide for a comparable population and time frame.

**Figure 3:**
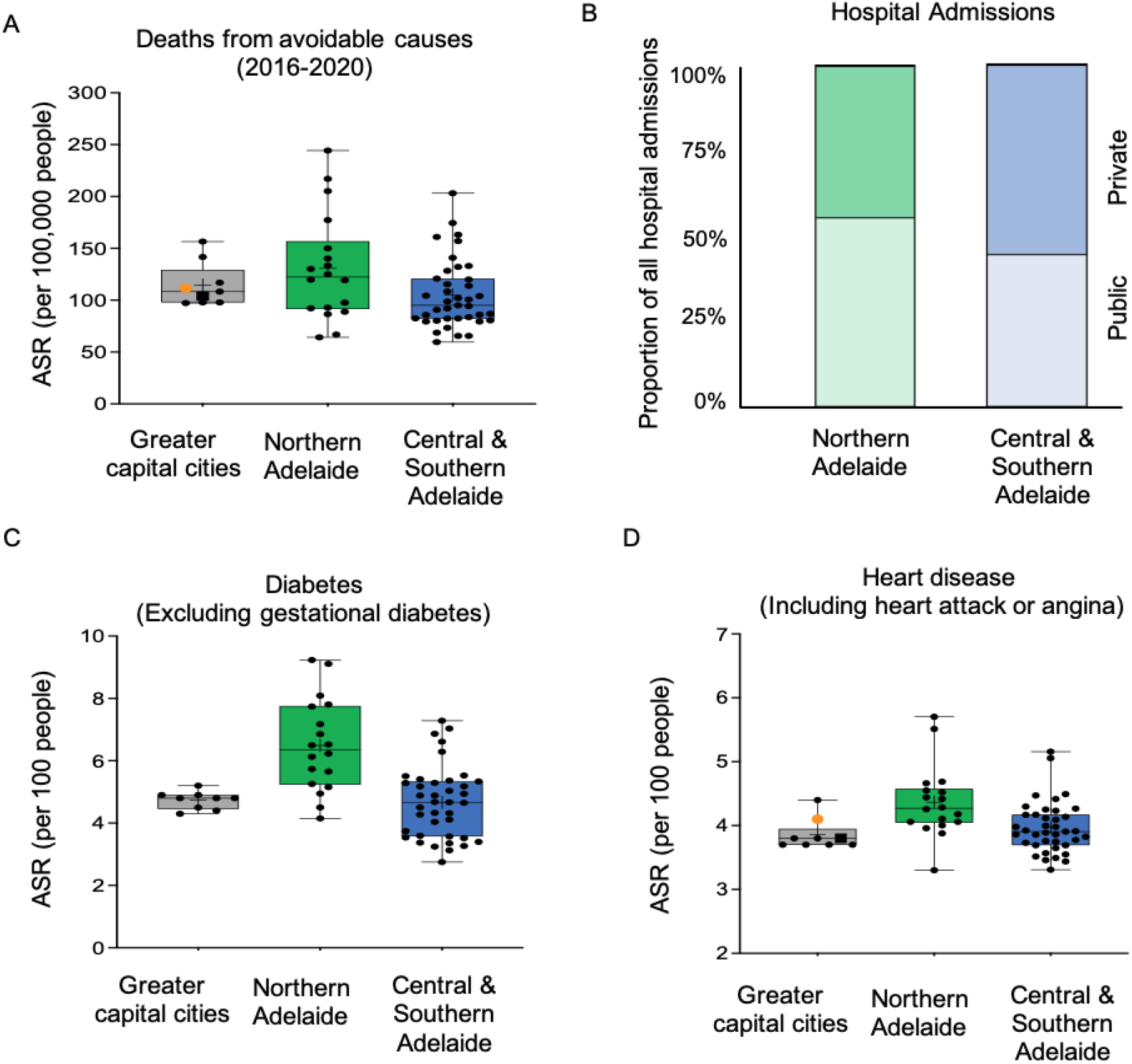
Profile of long-term health conditions and health measures. A) Deaths from avoidable causes (ASR per 100,000 people); B) proportion of private and public hospital admissions. ASR per 100 people with C) diabetes and D) heart-disease. Greater capital cities in Australia (grey ∼ Australian GCC = black square, greater Adelaide = orange circle), northern Adelaide region (green) and combined central and southern Adelaide regions (blue). Box and whisker plots, show the median, first, and third quartiles calculated for each group, with whisker boundaries representing the minimum and maximum values. All data from the 2021 census unless specified otherwise.

When considering the age standardised rate per 100,000 people the total admissions to all hospitals (excluding same-day admissions for renal dialysis) is reasonably consistent across the greater capital cities (Supplementary Table 3). In the northern Adelaide region 55% of all hospital admissions are to public hospitals, 22% greater the combined central and southern regions of Adelaide (Figure 3B).

The number of people self-reporting one or more or three or more long-term health conditions is higher in the northern Adelaide region compared to the combined central and southern regions of Adelaide (Supplementary Figure 4). These health conditions include diabetes (Figure 3C), heart disease (Figure 3D), kidney disease and lung conditions (Supplementary Figure 4). This equates to 27,170 people in the northern Adelaide region living with diabetes, 38% more than the relative rate in the central and southern regions of greater Adelaide for a comparable population (based on 2021 census).

### Risk Factors

A prevalent health condition in the northern Adelaide population is obesity (Table 2). In greater capital cities of Australia, the modelled estimates (2017-2018) for people over 18 years of age indicate that less than 30 in every 100 people (ASR) are obese. Greater Adelaide has one of the highest levels of people with obesity (32%), higher again in the northern Adelaide region at almost 37%. We demonstrate correlations between the higher numbers of people with obesity and the lower number of people with adequate fruit intake per day (Figure 4A), higher proportion of current smokers (Figure 4B) and higher numbers of people with low, very low or no levels of exercise (Figure 4C).

**Figure 4:**
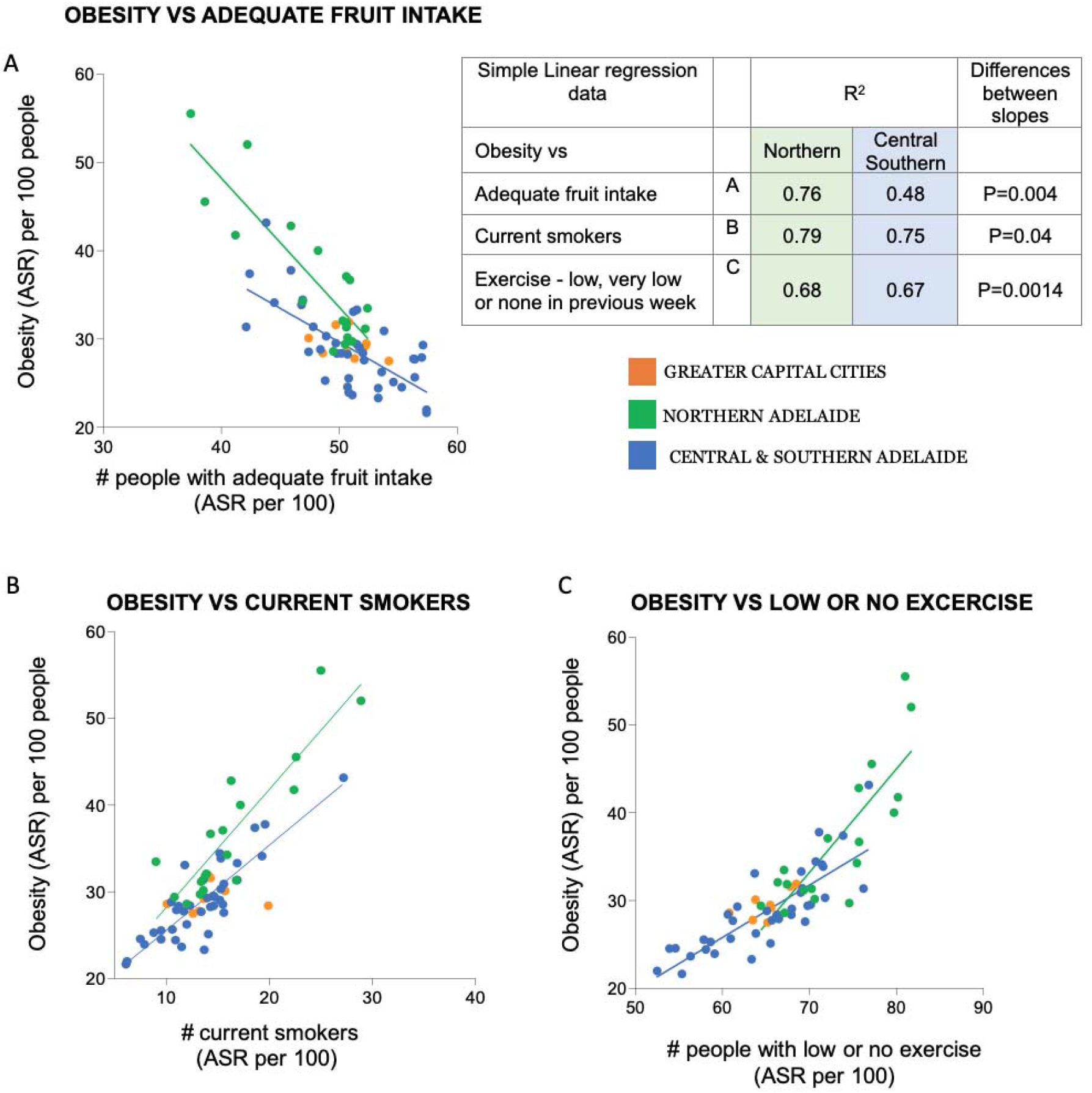
Association of the prevalence of obesity with risk factors in the northern Adelaide region. The number of people who were obese were compared to; A) the number of people with adequate levels of fruit intake; B) the number of people who were current smokers; C) the number of people who undertook low, very low or no exercise in previous week (exercise undertaken for fitness, sport or recreation in the week prior to being interviewed - level of exercise calculated ‘Duration of exercise (minutes) x Intensity factor (walking for fitness = 3.5, moderate = 5, vigorous = 7.5): low, very low or no exercise refers to scores of less than 800). All data were in people aged 18 yrs and over from modelled estimates across 2017-2018, with age standardised rate (ASR) per 100 people. Greater capital cities in Australia (orange) northern Adelaide region (green) and combined central and southern Adelaide regions (blue). Linear regression analysis for northern vs central and southern Adelaide regions is shown in table (top right panel).

## Discussion

Compared to other regions within greater Adelaide and greater capital cities nationally almost 80% of the northern Adelaide population resides in a local government area with an IRSD score indicating high relative disadvantage. The relative disadvantage of this northern Adelaide population is confronting. This community level disadvantage also occurs within other Australian capital cities^8^. The profile of social determinants of health indicates a community with a high percentage of people living on constrained incomes, lower levels of achieved education and a high percentage of families dependent on welfare. There are also high proportions of premature mortality and death from avoidable causes in this northern Adelaide population, including long-term health conditions such as diabetes. This disadvantaged community has higher proportions of people reporting factors contributing to poor health outcomes, including obesity. We demonstrate that the higher proportion of people with obesity in the north of Adelaide correlates with different risk factors impacting health outcomes including low numbers of people with adequate fruit intake and higher proportions of people who smoke and undertake low or no levels of exercise. Profiling the health and sociodemographic factors in this community provides key areas for health planners to focus efforts in creating and leading interventions aimed at improving health outcomes.

The profile of the northern Adelaide region captured in this study indicates a community with a high percentage of working age people receiving unemployment benefits and limited financial security. Protracted and long-lasting periods of parental unemployment is linked to long term health problems in their children ^9^ leading to an increased likelihood of inter-generational disadvantage. Without intervention it is likely that the social disadvantage and related health burden may continue to worsen for parts of this community. An example of this burden is shown by the study profile in the northern Adelaide community with a particularly prevalent incidence of obesity. Limited healthy food choices and opportunities for physical activity are known to contribute to the prevalence of obesity ^10-11^. A recent Australian study shows that most groceries purchased by low socioeconomic households between 2015 and 2019 consisted of ultra-processed foods associated that increase risks of obesity ^12^. This highlights the high priority public health need for education and access to healthy food choices. This is particularly pertinent given the current cost-of-living crisis which has already been shown to increase the rates of food and housing insecurity, affecting and exacerbating the disadvantaged disproportionally ^13-14^.

By profiling the social determinants of health and corresponding health status of the northern Adelaide community, we sought to better understand the scale of health needs for this population. Interestingly, the northern Adelaide community had a higher proportion of people under the age of 44 years compared to other regions of greater Adelaide. Despite the higher proportion of younger people in the population we observed higher proportions of long-term health conditions, premature deaths and deaths from avoidable causes as well as a higher proportion of admissions to public hospitals. There is a growing appreciation that inequity in health service provision and accessibility are linked to socioeconomic disadvantage and have been demonstrated recently in the Australian context for people with disabilities ^15^, people experiencing long term unemployment ^16^ and patients experiencing barriers to accessing digital health solutions offered via selected hospital clinics ^17^.

There are recent calls for targeted investment in vulnerable population interventions that may in turn provide necessary improvements to overall public health ^18^. The profile we provide in this study demonstrates the need for universal and targeted interventions to address the root cause of social disadvantage, focused on education, employment, and health literacy. Given the demonstrated dependence of this population on the public health system coupled with the underlying health conditions in this community, we contend that there is a demonstrated need in the short to medium term to ensure that there is adequate, locally based public infrastructure to meet the health care needs of this disadvantaged population, and that resourcing will need to be greater than that required in less disadvantaged areas. Despite the growing body of research by health care services on social care initiatives ^19^, there remains a clear need for robust research to establish the impact of treating an individual’s disease without adequately addressing underlying social disadvantage. At the coalface, clinicians and health workers have an important role to play in understanding how the social determinants of health, may impact patient care.

## Data Availability

We analysed data from the Australian Bureau of Statistics (ABS), using the Social Health Atlas of Australia: 2021 Census and data by population health areas (PHA), updated throughout 2023. This data is publicly available via https://phidu.torrens.edu.au

https://phidu.torrens.edu.au

## Data Sharing Statement

This article includes no original data.

## Conflict of interest

The authors declare no conflict of interest.

### List of Abbreviations

ASR: Age-standardised rate
CALHN: Central Adelaide local health network
LGA: local government area
LHN: local health network
NALHN: Northern Adelaide local health network
PHA: Public Health Area
SALHN: Southern Adelaide local health network

## List of URLs and online resources

Adelaide PHN https://adelaidephn.com.au

PHIDU https://phidu.torrens.edu.au

