## Supplemental Table 1 -3 for "Profiling health and socioeconomic disadvantage in the northern Adelaide Local Health Network population"

| **Supplementary Table 1 – LHN boundaries and PHA catchment for Greater Adelaide local Health Networks.** | | | | | | | |
| --- | --- | --- | --- | --- | --- | --- | --- |
| **NALHN** |  |  | **CALHN** |  |  | **SALHN** |  |
| **LGA code** | **LGA Name** |  | **LGA code** | **LGA Name** |  | **LGA code** | **LGA Name** |
| 45680 | Playford |  | 40070 | Adelaide City |  | 42600 | Holfast Bay |
| 45890 | Port Adelaide |  | 40700 | Burnside |  | 44060 | Marion |
| 47140 | Salsibury |  | 40910 | Campbeltown |  | 44060 | Marion |
| 47700 | Tea Tree Gully |  | 41060 | Charles Sturt |  | 44340 | Mitcham |
|  |  |  | 45290 | Norwood / Payneham -St Peters |  | 45340 | Onkaparinga |
|  |  |  | 45890 | Port Adelaide Enfield # |  |  |  |
|  |  |  | 46510 | Prospect |  |  |  |
|  |  |  | 47980 | Unley |  |  |  |
|  |  |  | 48260 | Walkerville |  |  |  |
|  |  |  | 48410 | West Torrens |  |  |  |
| **PHA code** | **PHA Name** |  | **PHA code** | **PHA Name** |  | **PHA code** | **PHA Name** |
| 40019 | Playford - West |  | 40000 | Adelaide |  | 40037 | Brighton (SA)/ Glenelg (SA) |
| 40020 | Davoren Park |  | 40001 | North Adelaide |  | 40038 | Edwardstown/ Morphettville |
| 40021 | Elizabeth East |  | 40006 | Burnside - Wattle Park |  | 40039 | Marion - South |
| 40022 | Elizabeth/ Smithfield - Elizabeth North |  | 40007 | Glenside - Beaumont/ Toorak Gardens |  | 40040 | Mitchell Park/ Warradale |
| 40023 | One Tree Hill |  | 40008 | Athelstone |  | 40041 | Belair/ Bellevue Heights/ Blackwood |
| 40024 | Enfield - Blair Athol |  | 40009 | Paradise - Newton |  | 40042 | Colonel Light Gardens/ Mitcham |
| 40025 | Northgate - Northfield/ Valley View - Gilles Plains |  | 40010 | Rostrevor - Magill |  | 40043 | Panorama |
| 40026 | Windsor Gardens |  | 40011 | Norwood (SA)/ St Peters - Marden |  | 40044 | Aberfoyle Park/ Coromandel Valley/ Flagstaff Hill |
| 40027 | Dry Creek - North/ Pooraka |  | 40012 | Payneham - Felixstow |  | 40045 | Aldinga |
| 40028 | Ingle Farm |  | 40013 | Nailsworth - Broadview/ Prospect/ Walkerville |  | 40046 | Christie Downs/ Hackham West - Huntfield Heights |
| 40029 | Para Hills/ Salisbury East |  | 40014 | Goodwood - Millswood |  | 40047 | Christies Beach/ Lonsdale |
| 40030 | Parafield/ Parafield Gardens/ Paralowie |  | 40015 | Unley - Parkside |  | 40048 | Clarendon/ McLaren Vale/ Willunga |
| 40031 | Salisbury/ Salisbury North |  | 40034 | Hope Valley - Modbury | 3% | 40049 | Hackham - Onkaparinga Hills/ Seaford |
| 40032 | Golden Grove/ Greenwith |  | 40053 | Beverley/ Hindmarsh - Brompton |  | 40050 | Happy Valley/ Happy Valley Reservoir/ Woodcroft |
| 40033 | Highbury - Dernancourt |  | 40054 | Flinders Park/ Seaton - Grange |  | 40051 | Morphett Vale - East/ Morphett Vale - West |
| 40034 | Hope Valley - Modbury | 97% | 40055 | Henley Beach |  | 40052 | Reynella |
| 40035 | Modbury Heights/ Redwood Park |  | 40056 | Charles Sturt - North West |  |  |  |
| 40036 | St Agnes - Ridgehaven |  | 40057 | West Lakes |  |  |  |
|  |  |  | 40058 | Dry Creek - South/ Port Adelaide/ The Parks |  |  |  |
| Where a PHA is split between a local health network boundary, the data were allocated to the PHA with the greater percentage footprint. | | | | | | | |

| **Supplementary Table 2: Country of origin of the population within greater capital cities and greater Adelaide regions** | | | | | | | | | | | |  |
| --- | --- | --- | --- | --- | --- | --- | --- | --- | --- | --- | --- | --- |
| **2021 Census** | **Northern** | Central + Southern | **Greater Adelaide *** |  | **Australia** (Greater CC^1^) | Greater Sydney | Greater Melbourne | Greater Brisbane | Greater Perth | Greater Hobart | Greater Darwin | Greater Canberra |
| Population (Total) | 417,994 | 851,826 | 1,387,290 |  | 17,020,559 | 5,231,147 | 4,917,750 | 2,526,238 | 2,116,647 | 247,086 | 139,902 | 454,499 |
| Australian born | 64.4 | 68.7 | 68.7 |  | 61.3 | 56.8 | 59.9 | 68.3 | 59.5 | 76.6 | 63.8 | 67.5 |
| Aboriginal Population | 2.4 | 1.3 | 1.7 |  | 1.8 | 1.7 | 0.7 | 3.0 | 2.0 | 4.5 | 10.4 | 2.0 |
| Born: English speaking countries | 8.2 | **8.7** | 8.4 |  | 8.5 | 6.9 | 6.3 | 10.8 | 15.5 | 5.9 | 6.2 | 6.3 |
| Born: non-English speaking countries | 21.3 | 19.1 | 19.2 |  | 25.8 | 31.8 | 29.4 | 12.9 | 20.5 | 13.0 | 21.6 | 22.4 |
| > 5yrs resident - AUS | 16.8 | 14.4 | 14.7 |  | 20.7 | 25.7 | 23.7 | 12.9 | 16.8 | 9.2 | 16.1 | 17.6 |
| < 5yrs resident - AUS | 3.9 | 4.2 | 3.9 |  | 4.4 | 5.4 | 5.0 | 3.0 | 2.9 | 3.5 | 4.9 | 4.3 |
| All data in table is as a % of Total population in greater capital cities (data in first row)  ^1^ GCC = Greater capital cities.  * Numbers of people self-reporting in Greater Adelaide Area are higher than the aggregate values of Southern +Central +Northern due to a small number of Public Health Areas that are captured in the Greater Adelaide region that fall outside the catchment for the three metropolitan Local Health Networks within Adelaide. | | | | | | | | | | | | |

|  | Supplementary Table 3: Prevalence of long-term health conditions and health measures in the greater capital cities and greater Adelaide regions | | | | | | | | | | | | |
| --- | --- | --- | --- | --- | --- | --- | --- | --- | --- | --- | --- | --- | --- |
|  | | **Northern**  **Adelaide** | Central + Southern | Greater Adelaide* |  | Australia (Greater CC^1^) | Greater Sydney | Greater Melbourne | Greater Brisbane | Greater Perth | Greater Hobart | Greater Darwin | Greater Canberra |
|  | **NUMBER AND TYPE OF LONG-TERM HEALTH CONDITIONS (2021)** | | | | | | | | | | | | |
| - 1 or more | | 31.1 | 28.2 | 29.3 |  | 26.7 | 24.4 | 26.5 | 29.9 | 26.7 | 31.2 | 23.1 | 30.0 |
| - 3 or more | | 4.1 | 3.0 | 3.3 |  | 2.8 | 2.5 | 2.7 | 3.5 | 2.6 | 3.5 | 2.5 | 2.9 |
| Diabetes ^2^ | | 6.5 | 4.7 | 5.2 |  | 4.8 | 4.9 | 4.8 | 4.9 | 4.5 | 4.3 | 4.8 | 4.4 |
| Heart Disease ^3^ | | 4.4 | 4.0 | 4.1 |  | 3.8 | 3.7 | 3.7 | 4.4 | 3.8 | 3.8 | 3.7 | 3.7 |
| Kidney Disease | | 0.94 | 0.75 | 0.8 |  | 0.9 | 0.9 | 0.9 | 1.0 | 0.8 | 0.9 | 1.0 | 1.1 |
| Lung Conditions ^4^ | | 2.1 | 1.6 | 1.7 |  | 1.5 | 1.3 | 1.3 | 2.1 | 1.6 | 1.9 | 1.8 | 1.6 |
|  | **MEASURES OF HEALTH (2016 to 2020)** | | | | | | | | | | | | |
| Median age at death- persons ^5^ | | 79 | 83 | 83 |  | 82 | 82 | 82 | 80 | 81 | 81 | 70 | 81 |
| Premature mortality – Total deaths 0 -74 years ^6^ | | 270.8 | 223.6 | 235.6 |  | 213.1 | 205.1 | 200.7 | 232.8 | 213.7 | 270.2 | 281.9 | 199.4 |
| Death from avoidable causes, 0-74 years ^5^ | | 130.6 | 105.2 | 111.4 |  | 104.0 | 97.2 | 97.9 | 116.9 | 108.3 | 141.7 | 156.6 | 97.9 |
| **HOSPITAL ADMISSIONS (2018-2019)** | | | | | | | | | | | | | |
| All hospitals ^7^ | | 35,413 | 37,319 | 36,892 |  | 37,178 | 35,399 | 39,698 | NA | 37,660 | NA | NA | NA |
| Private hospitals ^7^ | | 15,647 | 20,598 | 19,002 |  | 17,878.2 | 17,881 | 17,621 | NA | 17,682 | NA | NA | NA |
| Public hospitals ^7^ | | 19,485 | 16,543 | 17,807 |  | 20,527.7 | 17,442 | 21,917 | 26,026 | 19,872 | 18,229 | 33,030 | 20,706 |
|  | ^1^ GCC = Greater capital cities.  Number and type of long-term health conditions - All data presented as Age Standardised Rate per 100 people self-reporting having a long-term health condition as part of the 2021 census (Australian Bureau of Statistics). ^2^ Excluding gestational diabetes; ^3^ Including heart attack or angina; ^4^ Including chronic obstructive pulmonary disease or emphysema.  Measures of health - ^5^ Average value of the collective of individual Public Health Area Median age of death; ^6^ Average annual Age standardised rate per 100,000 **(**2016-2020).  Hospital admissions exclude same-day admissions for renal dialysis. ^7^ Average Age standardised rate per 100,000 people (rounded to nearest whole number). | | | | | | | | | | | | |
