## Supplementary Figures 1-4 for "Profiling health and socioeconomic disadvantage in the northern Adelaide Local Health Network population"

**Supplementary data and figures**

**The northern Adelaide region is a region of disadvantage.**

The distribution of the IRSD scores for the individual public health areas (PHA) are shown as individual points in the graph in Supplementary Figure 1 and highlight that most PHAs within this region are below the national average of 1000 with several showing very low IRSD scores indicative of severe disadvantage.

Northern Adelaide

Central & Southern Adelaide

Greater capital cities

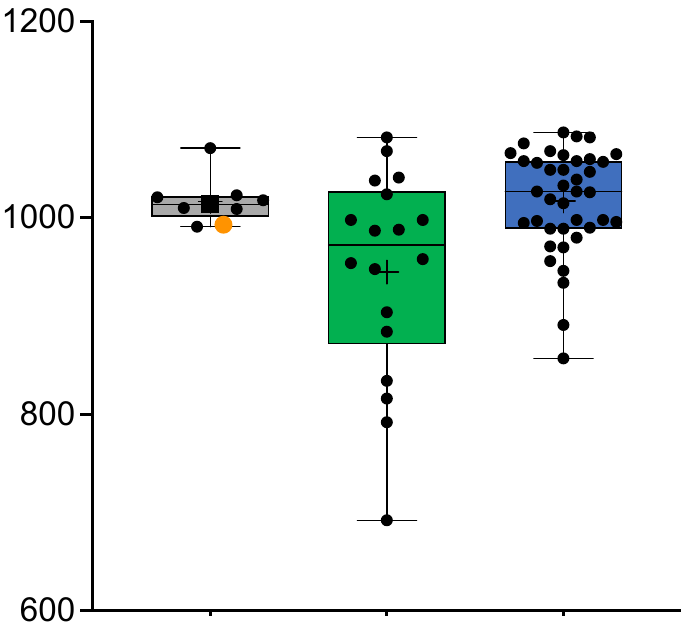

**Supplementary Figure 1: IRSD scores in greater Adelaide is being driven by the relative disadvantage in the northern Adelaide region**. IRSD scores for aggregated PHAs within LGA areas for the greater Adelaide region where the northern region (green), and central and southern Adelaide regions (blue). Greater capital cities index scores for Australia (grey bar; Australian GCC as a black square, greater Adelaide as an orange dot). Data presented as box and whisker plots, with the median, first, and third quartiles calculated for each group, and whisker boundaries representing the minimum and maximum values. Red dotted line indicates the national average IRSD score.

**Supplementary Figure 2: Age distribution in greater capital cities of Australia**.

Broad age distribution for the Greater capital cities (grey bars) (percentage of Total persons). Data from 2020 estimated resident population, released in 2021.

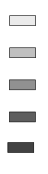

0-14 yrs

15-24 yrs

25-44 yrs

45-64 yrs

>65 yrs

Australia	Sydney	Melbourne	Brisbane	Adelaide	Perth	Hobart	Darwin	Canberra

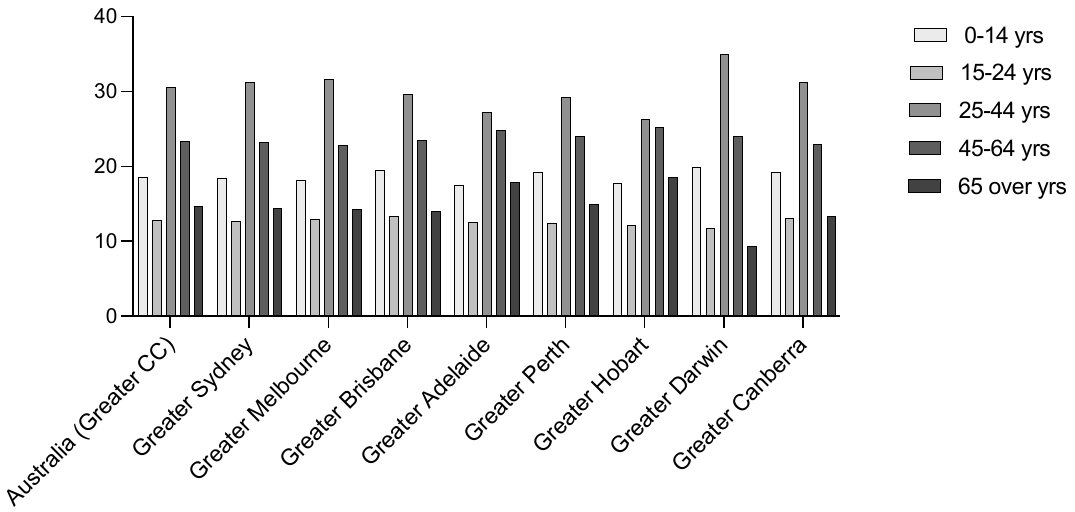

% of Total Persons (per broad age group)

Greater capital cities

NORTHERN ADELAIDE

CENTRAL & SOUTHERN ADELAIDE

GREATER CAPITAL CITIES

% people 16-64 yrs

Unemployment benefits

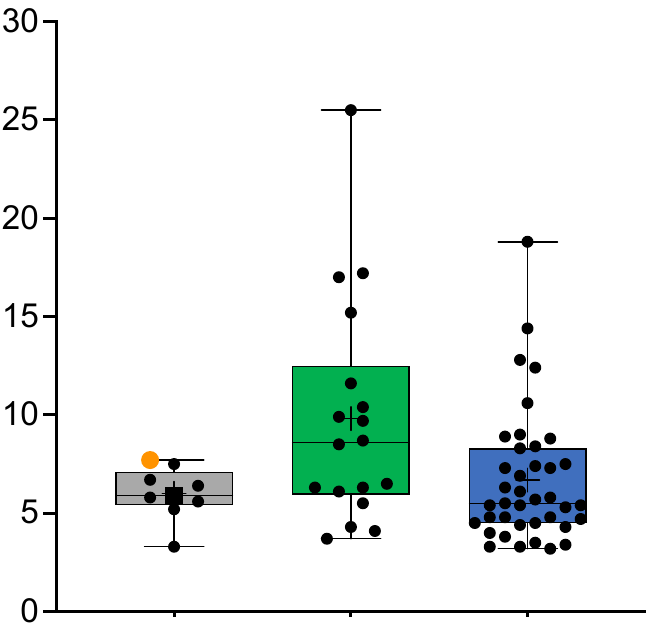

B

A

Schooling less than Yr 10

Age standardised rate per 100 people

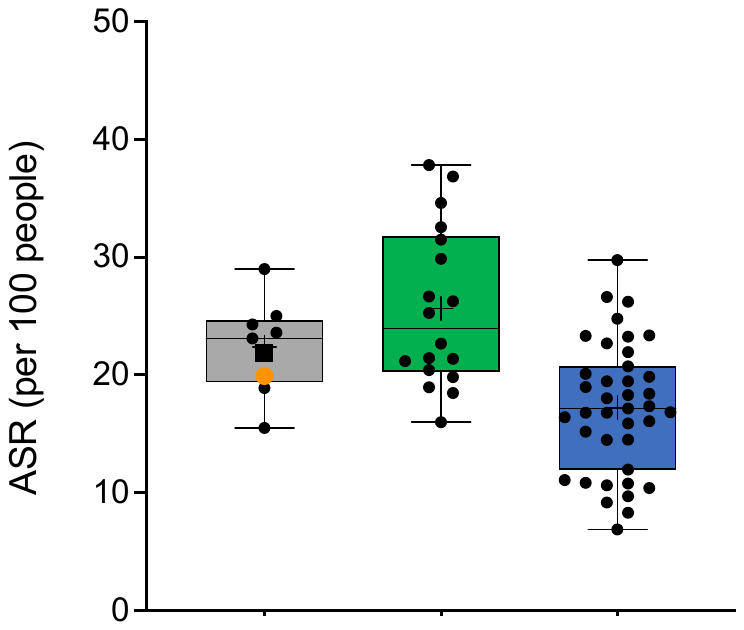

% people 65 yrs and older

Age pension

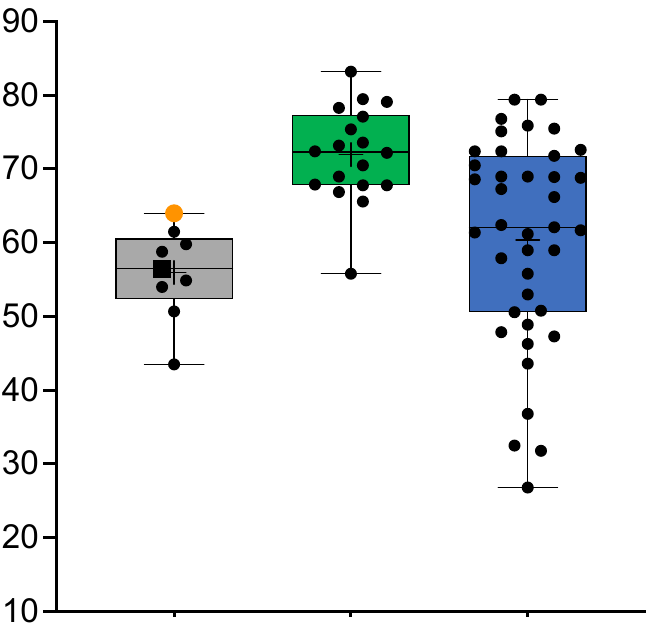

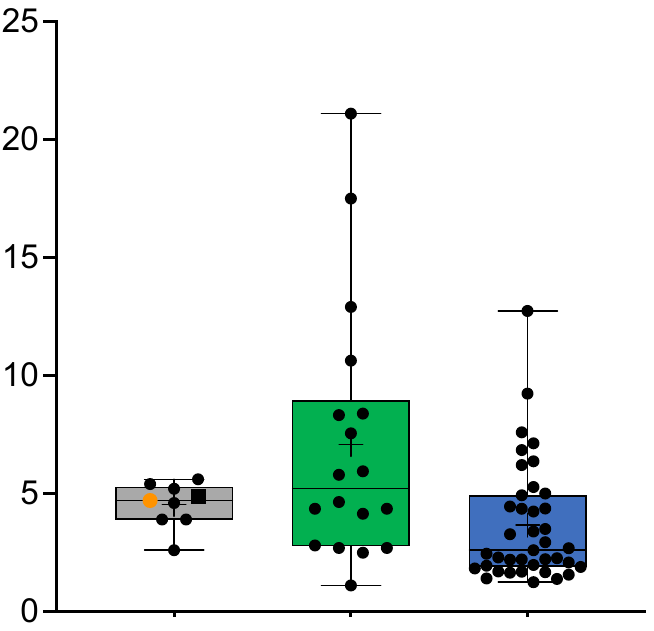

Low-income welfare dependent families with children

% Total families

D

C

F

E

% social housing

(Rented dwellings)

Social housing (rented)

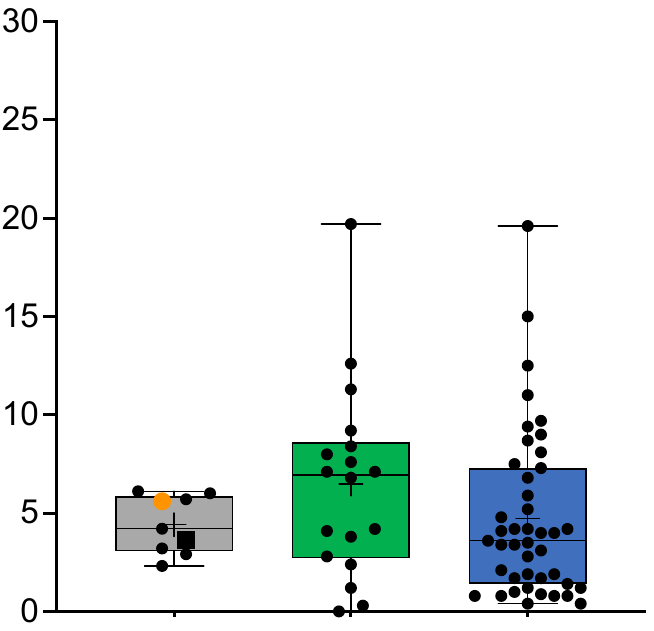

% households in dwellings receiving rent assistance

Rent assistance

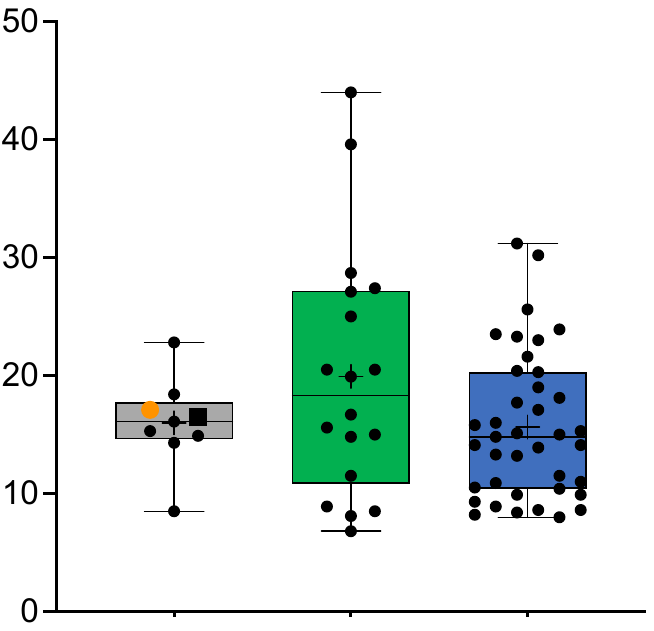

**Supplementary Figure 3: Social factors contributing to disadvantage in northern Adelaide.**

A) Number of people with schooling less than year 10 (ASR per 100 people); B) percentage of people aged 16-64 yrs receiving unemployment (Newstart or youth allowance) benefits; C) percentage of people over the age of 65 years receiving the age pension; D) proportion of low-income welfare dependent families with children, E) proportion of social housing (rented) and F) proportion of households receiving rent assistance. Greater capital cities in Australia (grey ~ Australian GCC = black square, greater Adelaide = orange circle), northern Adelaide region (green) and combined central and southern Adelaide regions (blue). Box and whisker plots, show the median, first, and third quartiles calculated for each group, with whisker boundaries representing the minimum and maximum values.

B

A

ASR (per 100 people)

One or more long-term health conditions

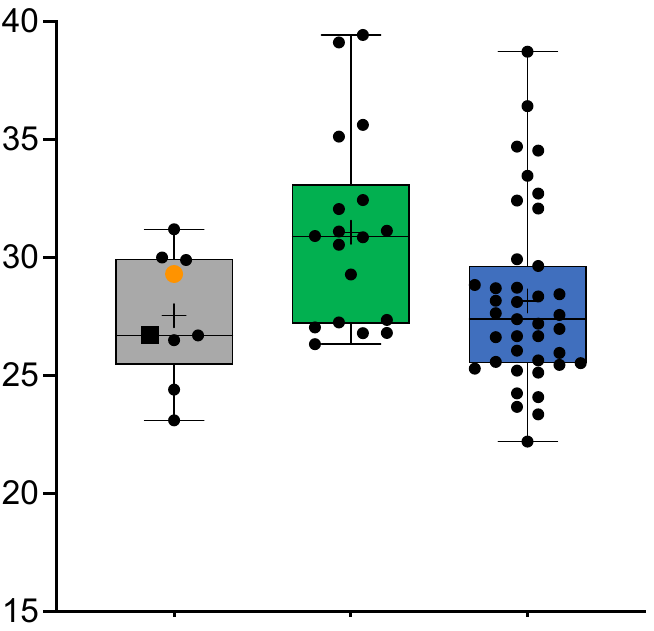

Northern Adelaide

Central & Southern Adelaide

Greater capital cities

ASR (per 100 people)

Three or more long-term health conditions

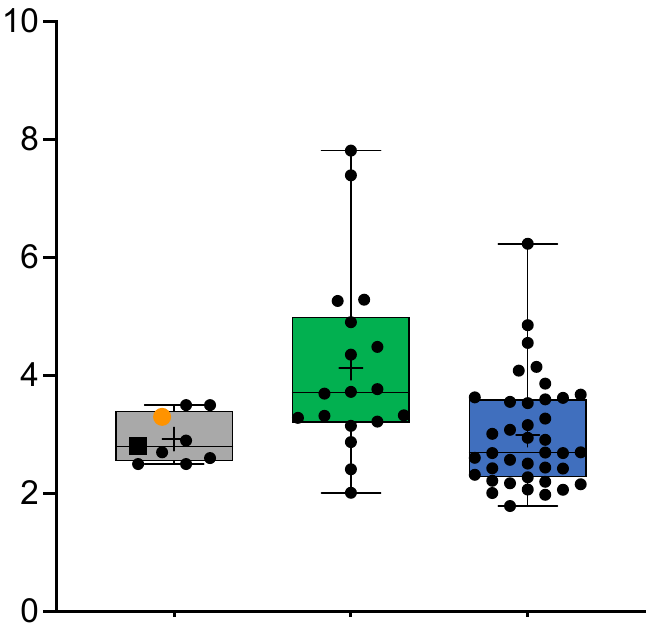

Northern Adelaide

Central & Southern Adelaide

Greater capital cities

D

C

ASR (per 100 people)

Lung conditions

(Including COPD or emphysema)

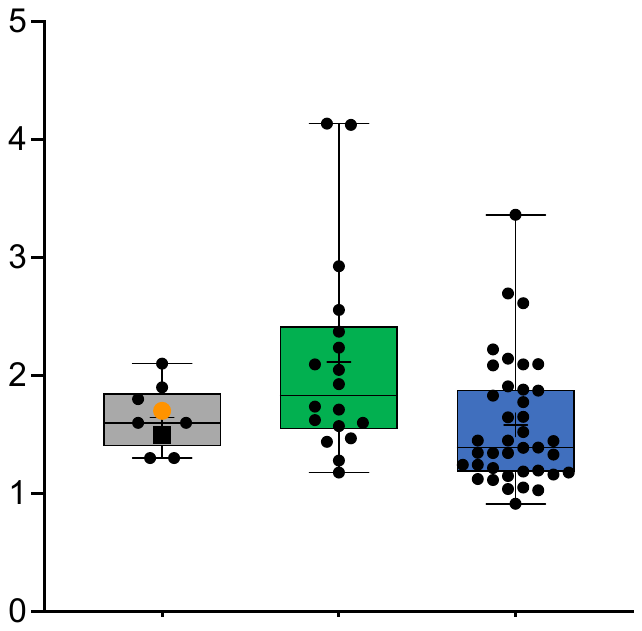

Northern Adelaide

Central & Southern Adelaide

Greater capital cities

ASR (per 100 people)

Kidney disease

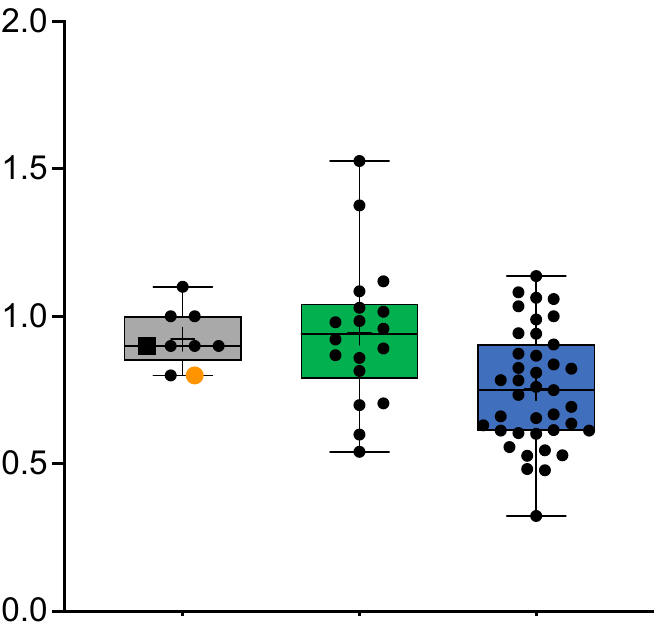

Northern Adelaide

Greater capital cities

Central & Southern Adelaide

**Supplementary Figure 4: Long-term health conditions and health measures.**

A) Number of people with one or more and B) three or more long-term health disorders (self-reported) (ASR per 100 people). ASR per 100 people with C) kidney-disease and D) lung conditions as a self-reported long term health condition. Greater capital cities in Australia (grey ~ Australian GCC = black square, greater Adelaide = orange circle), northern Adelaide region (green) and combined central and southern Adelaide regions (blue). Box and whisker plots, show the median, first, and third quartiles calculated for each group, with whisker boundaries representing the minimum and maximum values.
