## Supplementary File 1 for "Profiling health and socioeconomic disadvantage in the northern Adelaide Local Health Network population": Supplementary File 1.pdf

### National Healthcare Agreement: PI 16–Potentially avoidable deaths, 2020

#### Identifying and definitional attributes

**Metadata item type:** Indicator

**Indicator type:** Progress measure

**Short name:** PI 16–Potentially avoidable deaths, 2020

**METEOR identifier:** 716490

**Registration status:**

- Health, Standard 13/03/2020

**Description:** Deaths from conditions that are potentially preventable through individualised care and/or treatable through existing primary or hospital care.

**Indicator set:** National Healthcare Agreement (2020)
Health, Standard 13/03/2020

**Outcome area:** Primary and Community Health
Health, Standard 07/07/2010

#### Collection and usage attributes

**Population group age to:** 74 years

**Computation description:** Deaths are defined as avoidable in the context of the present health system.

International Classification of Diseases (ICD-10, 2016 version) codes in scope are as specified below:

**Table 1: Potentially avoidable deaths International Classification of Disease (ICD-10) Codes**

| Cause of death groups | ICD-10 Codes | Limits (age, sex) |
|---|---|---|
| **Infections** | | |
| Selected invasive infections | A38–A41, A46, A48.1, G00, G03, J02.0, J13–J16, J18, L03 | |
| Viral pneumonia and influenza | J10–J12 | |
| HIV/AIDS | B20–B24 | |
| **Cancer** | | |
| Cancer of the colon, sigmoid, rectum and anus | C18–C21, C26.0 | |
| Skin | C43, C44 | |
| Breast | C50 | Female |
| Cervix | C53 | |
| Prostate | C61 | |
| Kidney | C64 | |
| Thyroid | C73 | |
| Hodgkin's disease | C81 | |
| Acute lymphoid leukaemia/Acute lymphoblastic leukaemia | C91.0 | 0-44 years |

| | |
|---|---|
| **Diabetes** | E10–E14 |
| **Diseases of the circulatory system** | |
| Rheumatic and other valvular heart disease | I00–I09, I33–I37 |
| Hypertensive heart and renal disease | I10–I13 |
| Ischaemic heart disease | I20–I25 |
| Cerebrovascular diseases | I60–I69 |
| Heart failure | I50, I51.1, I51.2, I51.4, I51.5 |
| Pulmonary embolism | I26 |
| **Diseases of the genitourinary system** | |
| Renal failure | N17–N19 |
| **Diseases of the respiratory system** | |
| COPD | J40–J44 |
| Asthma | J45, J46 |
| **Diseases of the digestive system** | |
| Peptic ulcer disease | K25–K27 |
| **Maternal & infant causes** | |
| Complications of the perinatal period | P00–P96 |
| **Other conditions** | |
| Complications of pregnancy, labour or the puerperium | O00–O99 |
| **Selected external causes of morbidity and mortality** | |
| Falls | W00–W19 |
| Fires, burns | X00–X09 |
| Suicide and self-inflicted injuries | X60–X84, Y87.0 |
| Misadventures to patients during surgical and medical care | Y60–Y69 |
| Medical devices associated with adverse incidents in diagnostic and therapeutic use | Y70–Y82 |
| Surgical and other medical procedures as the cause of abnormal reaction of the patient, or of later complication, without mention of misadventure at the time of the procedure | Y83, Y84 |
| **Other external causes of morbidity and mortality** | |
| Transport accidents | V01–V99 |
| Exposure to inanimate mechanical forces | W20–W49 |
| Exposure to animate mechanical forces | W50–W64 |
| Accidental drowning and submersion | W65–W74 |
| Other accidental threats to breathing | W75–W84 |
| Exposure to electric current, radiation and extreme ambient air temperature and pressure | W85–W99 |
| Contact with heat and hot substances | X10–X19 |
| Contact with venomous animals and plants | X20–X29 |
| Exposure to forces of nature | X30–X39 |

| | |
|---|---|
| Accidental poisoning by and exposure to noxious substances | X40–X49 |
| Overexertion, travel and privation | X50–X57 |
| Accidental exposure to other and unspecified factors | X58,X59 |
| Assault | X85–Y09 |
| Event of undetermined intent | Y10–Y34 |
| Legal interventions and operations of war | Y35, Y36 |
| Drugs, medicaments and biological substances causing adverse effects in therapeutic use | Y40–Y59 |
| Sequelae of external causes of morbidity and mortality | Y85, Y86, Y87.1–Y89 |

Rates are directly age-standardised to the 2001 Australian population.

Variability bands are to be calculated for single-year rates using the method below.

Presented per 100,000 population.

**Computation:** Number

100,000 x (Numerator ÷ Denominator)

Variability bands are to be calculated for single-year rates using the following method for estimating 95% confidence intervals:

*Age-standardised rate*

$$CI\ (ASR)_{95\%} = \mathrm{ASR}\ \pm 1.96\ \times \sqrt{\sum_{i=1}^{i} \frac{w_i^2\, d_i}{n_i^2}}$$

Where $w_i$= the proportion of the standard population in age group *i*

$d_i$=the number of deaths in age group *i*

$n_i$=the number of people in the population in age group *i*

**Numerator:** Number of deaths of persons aged less than 75 categorised as potentially avoidable

**Numerator data elements:**

**Data Element / Data Set**

**Data Element**

Person—age

**Data Source**

ABS Causes of Death Collection

**Guide for use**

Data source type: Administrative by-product data

**Data Element / Data Set**

**Data Element**

Person—underlying cause of death, code (ICD-10 2016 version) ANN{.N}

**Data Source**

ABS Causes of Death Collection

**Guide for use**

Data source type: Administrative by-product data

**Denominator:** Population aged less than 75

**Denominator data elements:**

**Data Element / Data Set**

**Data Element**

Person—projected Indigenous population of Australia, total people N[N(7)]

**Data Source**

ABS Indigenous estimates and projections (2016 Census-based)

**Guide for use**

Data source type: Census-based plus administrative by-product data

**Data Element / Data Set**

**Data Element**

Person—age, total years N[NN]

**Data Source**

ABS Estimated resident population (2016 Census-based)

**Guide for use**

Data source type: Census-based plus administrative by-product data

**Data Element / Data Set**

**Data Element**

Person—age, total years N[NN]

**Data Source**

ABS Indigenous estimates and projections (2016 Census-based)

**Guide for use**

Data source type: Census-based plus administrative by-product data

**Data Element / Data Set**

**Data Element**

Person—estimated resident population of Australia, total people N[N(7)]

**Data Source**

ABS Estimated resident population (2016 Census-based)

**Guide for use**

Data source type: Census-based plus administrative by-product data

**Disaggregation:**

2015, 2016, 2017 (resupplied for revision to ABS cause of death data), 2018—State and territory.

2015, 2016, 2017 (updated for revision to ABS cause of death data), 2018—Nationally, by Indigenous status (not reported).

2011–2015, 2012–2016, 2013–2017 (updated for revision to ABS cause of death data), 2014–2018—State and territory, by Indigenous status.

Some disaggregations may result in numbers too small for publication. Disaggregation by Indigenous status will be based on data only from jurisdictions for which the quality of Indigenous identification is considered acceptable—New South Wales, Queensland, South Australia, Western Australia, Northern Territory.

**Disaggregation data elements:**

**Data Element / Data Set**

**Data Element**

Person—Indigenous status, code N

**Data Source**

ABS Causes of Death Collection

**Guide for use**

Data source type: Administrative by-product data

**Data Element / Data Set**

**Data Element**

Person—area of usual residence, statistical area level 2 (SA2) code (ASGS 2016) N(9)

**Data Source**

ABS Causes of Death Collection

**Guide for use**

Data source type: Administrative by-product data
Used for disaggregation by state/territory

**Data Element / Data Set**

**Data Element**

Person—underlying cause of death, code (ICD-10 2016 version) ANN{.N}

**Data Source**

ABS Causes of Death Collection

**Guide for use**

Data source type: Administrative by-product data

**Comments:**

Most recent data available for 2020 National Healthcare Agreement performance reporting:

- 2018 (Total population and Indigenous status at national level)
- Aggregated data 2014–2018 (Indigenous status)

A number of updates to the ICD-10 were applied to 2013 and subsequent years causes of death data. Details of the impact of these changes on the mortality data are described in ABS Implementation of Iris Software: Understanding Coding and Process Improvements.

2013 data are coded using ICD-10 (2013 version). 2014, 2015, 2016 and 2017 data are coded using ICD-10 (2015 version). 2018 data are coded using ICD-10 (2016 version).

Due to small number of Indigenous deaths reported each year, 5-year combined data will be reported for state and territory disaggregations.

Estimated Residential Population (ERP) data for the total population and the Indigenous population are sourced from ERP rebased after the 2016 Census.

Data by remoteness may be available, pending assessment of data quality.

#### Representational attributes

**Representation class:** Rate

**Data type:** Real

**Unit of measure:** Person

**Format:** NN[N].N

#### Indicator conceptual framework

**Framework and dimensions:** Deaths

#### Data source attributes

**Data sources:**

**Data Source**

ABS Indigenous estimates and projections (2016 Census-based)

**Frequency**

Periodic

**Data custodian**

Australian Bureau of Statistics

**Data Source**

ABS Causes of Death Collection

**Frequency**

Annual

**Quality statement**

ABS causes of death collection, QS

**Data custodian**

Australian Bureau of Statistics

**Data Source**

ABS Estimated resident population (2016 Census-based)

**Frequency**

Quarterly

**Data custodian**

Australian Bureau of Statistics

#### Accountability attributes

**Reporting requirements:** National Healthcare Agreement

**Organisation responsible for providing data:** Australian Bureau of Statistics (ABS).

**Further data development / collection required:** Specification: Minor work required, the measure needs minor work to meet the intention of the indicator.

#### Source and reference attributes

**Reference documents:** ABS (Australian Bureau of Statistics) 2015. Causes of Death, Australia, 2013. ABS cat.no. 3303.0. Canberra: ABS

#### Relational attributes

**Related metadata references:**

Supersedes National Healthcare Agreement: PI 16–Potentially avoidable deaths, 2019

- Health, Superseded 13/03/2020

Has been superseded by National Healthcare Agreement: PI 16–Potentially avoidable deaths, 2021

- Health, Standard 03/07/2020

See also Australian Health Performance Framework: PI 1.2.1–Rates of current daily smokers, 2019

- Health, Standard 09/04/2020

See also Australian Health Performance Framework: PI 1.2.1–Rates of current daily smokers, 2020

- Health, Standard 13/10/2021

See also Australian Health Performance Framework: PI 1.2.3–Levels of risky alcohol consumption, 2019

- Health, Standard 09/04/2020

See also Australian Health Performance Framework: PI 1.2.3–Levels of risky alcohol consumption, 2020

- Health, Standard 13/10/2021

See also Australian Health Performance Framework: PI 1.3.1–Prevalence of overweight and obesity, 2019

- Health, Standard 09/04/2020

See also Australian Health Performance Framework: PI 1.3.1–Prevalence of overweight and obesity, 2020

- Health, Standard 13/10/2021

See also Australian Health Performance Framework: PI 2.1.4–Selected potentially preventable hospitalisations, 2019

- Health, Standard 09/04/2020

See also Australian Health Performance Framework: PI 2.1.4–Selected potentially preventable hospitalisations, 2020

- Health, Standard 01/12/2020

See also Australian Health Performance Framework: PI 2.1.6–Potentially avoidable deaths, 2019

- Health, Standard 09/04/2020

See also Australian Health Performance Framework: PI 2.1.6–Potentially avoidable deaths, 2020

- Health, Standard 01/12/2020

See also National Healthcare Agreement: PI 03–Prevalence of overweight and obesity, 2020

- Health, Standard 13/03/2020

See also National Healthcare Agreement: PI 04–Rates of current daily smokers, 2020

- Health, Standard 13/03/2020

See also National Healthcare Agreement: PI 05–Levels of risky alcohol consumption, 2020

- Health, Standard 13/03/2020

See also National Healthcare Agreement: PI 06–Life expectancy, 2020

- Health, Standard 13/03/2020

See also National Healthcare Agreement: PI 07–Infant and young child mortality rate, 2020

- Health, Standard 13/03/2020

See also National Healthcare Agreement: PI 08–Major causes of death, 2020

- Health, Standard 13/03/2020

See also National Healthcare Agreement: PI 18–Selected potentially preventable hospitalisations, 2020

- Health, Standard 13/03/2020

See also National Healthcare Agreement: PI 23–Unplanned hospital readmission rates, 2020

- Health, Standard 13/03/2020

© Australian Institute of Health and Welfare 2023

This product, excluding the AIHW logo, Commonwealth Coat of Arms and any material owned by a third party or protected by a trademark, has been released under a Creative Commons BY 3.0 (CC BY 3.0) licence. Excluded material owned by third parties may include, for example, design and layout, images obtained under licence from third parties and signatures. We have made all reasonable efforts to identify and label material owned by third parties.

You may distribute, remix and build upon this work. However, you must attribute the AIHW as the copyright holder of the work in compliance with our attribution policy available at www.aihw.gov.au/copyright. The full terms and conditions of this licence are available at http://creativecommons.org/licenses/by3.0/au/.

Enquiries relating to copyright should be addressed to the Head of the Communications, Media and Marketing Unit, Australian Institute of Health and Welfare, GPO Box 570, Canberra ACT 2601.
